# Association of Cardiovascular Health and Genetic Risk for Left Ventricular Mass with Left Ventricular Structure, Function, and Incident Cardiovascular Outcomes

**DOI:** 10.64898/2026.01.07.26343640

**Authors:** Marcela Cuellar-Lobo, Musong Gao, Maria M. Ruda, Brenton Prescott, Vanessa Xanthakis, Emelia J. Benjamin, Susan Cheng, Ramachandran S. Vasan, Ching-Ti Liu, Connie W. Tsao

**Affiliations:** Cardiovascular Division, Department of Medicine, Beth Israel Deaconess Medical Center, Harvard Medical School, Boston, MA; Department of Biostatistics, Boston University School of Public Health, 801 Massachusetts Ave, Boston, MA; Cardiovascular Imaging Core Laboratory, Division of Cardiovascular Medicine, Brigham and Women’s Hospital, Harvard Medical School, Boston, MA; Section of Preventive Medicine and Epidemiology, Department of Medicine, Boston University Chobanian & Avedisian School of Medicine, Boston, MA; Framingham Heart Study, Framingham, MA; Section of Cardiovascular Medicine, Department of Medicine, Boston Medical Center and Boston University Chobanian & Avedisian School of Medicine, Boston, MA and Department of Epidemiology, Boston University School of Public Health; Department of Cardiology, Smidt Heart Institute, Cedars-Sinai Medical Center, Los Angeles, CA, USA; University of Texas School of Public Health and Departments of Medicine and Population Health Sciences, University of Texas Health Science Center, San Antonio, TX

**Keywords:** Life’s Essential 8, Genetic risk, Cardiovascular outcomes, cohort study, echocardiography

## Abstract

**Background:** Cardiovascular health (CVH) and genetic susceptibility to adverse left ventricular (LV) remodeling are each linked to cardiovascular diseases (CVD); however, their combined role remains unclear.

**Methods:** Framingham Heart Study participants [n=1,255 Offspring-Exam 8 (2005-2008), n=2,835 Generation 3-Exam 1 (2002-2005)] had assessment of modifiable risk factors comprising the American Heart Association’s Life’s Essential 8 (LE8) score with higher scores indicating better CVH health. A Polygenic Risk Score (PRS) for LV mass index (LVMI) was also calculated based on published PRS(PGS003427). We created 9 groups combining LE8 and PRS tertiles (high LE8+low PRS served as referent group reflecting optimal risk) and related these groups to presence of LVMI, LV ejection fraction (LVEF), and the ratio of the peak early diastolic mitral inflow velocity (E wave) to the peak early diastolic mitral annular velocity (e’ wave; E/e’; separate model for each outcome), and incident cardiovascular disease (CVD; a composite of coronary heart disease [CHD], heart failure, stroke, peripheral arterial disease). We applied linear mixed regression and Cox regression models to evaluate the relation of LE8-PRS groups with all outcomes mentioned above.

**Results:** Participants (56% women, mean age 47 years) had mean LE8 score 70 (SD=13), indicating intermediate CV health and normal LVMI and systolic function (LVMI 76±13 g/m², LVEF 65±5%, E/e’ 6 ±1.8 cm/s). Over 19 years of follow-up, composite CV events and CHD occurred in n=188 and n=99, respectively, of Offspring and n=83 and n=49 respectively, of Gen 3 participants. Compared to the referent group, individuals in the low LE8-high PRS group had high LVMI, E/e’, and over three-fold higher risks for CVD and CHD, with incidence rates of approximately 1.84 versus 4.06 per 1,000 person-year, respectively.

**Conclusion:** Lower LE8 scores (indicating worse CVH) combined with high genetic risk confer higher conjoint risks for adverse LV structure, function, and CVD development.

## INTRODUCTION

Cardiovascular disease (CVD) remains the leading cause of morbidity and mortality, responsible for nearly one-third of global deaths, imposing substantial healthcare costs, with modifiable risk factors and genetic predisposition playing central roles in its development ^1,2^. To quantify modifiable risk, the American Heart Association (AHA) introduced *Life’s Essential 8* (LE8), a comprehensive metric assessing cardiovascular health (CVH) across eight domains: four health behaviors (diet, physical activity, nicotine exposure, and sleep) and four health factors (body mass index, blood lipids, blood glucose, and blood pressure) ^3^. Each component is scored from 0 (poor health) to 100 (ideal health), and the total LE8 score is calculated as an unweighted average of the 8 component scores ^3^, with higher LE8 scores indicating better CVH. The LE8 score has been associated with lower risks of CVD and mortality ^4,5^; however, despite advances in prevention, suboptimal CVH in adults persists, demonstrated by mean LE8 scores well below optimal levels across diverse demographic groups in national surveys ^6,7^.

Parallel to modifiable risk factors, inherent genetic risk for traits has long been demonstrated. In the past decade, advances in genomics have led to the development of polygenic risk scores (PRS), which aggregate the influence of thousands of common genetic variants to estimate an individual’s inherited susceptibility to disease ^8–10^. PRS for given traits may be added to routine screening to identify high-risk individuals who may benefit from early interventions, including enhanced screening or more aggressive preventive measures, such as lifestyle modifications or medical therapy ^9^. One trait of adverse CV remodeling is left ventricular mass index (LVMI), elevations of which are strongly associated with risk of heart failure (HF), stroke, and coronary heart disease (CHD) in individuals without established CVD ^11^. Early identification of individuals at risk for elevated LVMI and incident CVD using PRS may enable primary prevention strategies ^11–13^.

Previous studies have demonstrated that the combination of modifiable CV risk factor levels with PRS of single traits such as hypertension or dyslipidemia jointly influence cardiovascular risk ^14,15^. Although both LE8 and PRS are strongly related to incident CVD, their associations with LVMI, a key intermediate phenotype, are less well characterized ^16^. LVMI reflects cumulative exposure to adverse health behaviors and genetic predisposition ^7^. Understanding these relationships may inform risk stratification, guide personalized prevention strategies, and identify individuals who would benefit most from early intervention. Furthermore, clarifying the role of LVMI in the pathway from LE8 and PRS to clinical events could enhance mechanistic insights and refine predictive models for CVD. We aimed to investigate these associations using the Framingham Heart Study, a large, well-phenotyped population-based cohort with detailed assessment of CV risk factors, genetic data, cardiac imaging, and longitudinal follow-up for CV outcomes.

## METHODS

### Study Participant and Design

We collected data from the Framingham Heart Study (FHS) Offspring cohort, Exam 8 (n= 2,809, 2005-2008) and Third Generation (Gen3) cohort, Exam 1 (n=4,080, 2002-2005), whose study designs have been previously described ^17,18^. During these examinations, participants underwent a comprehensive history and physical examination conducted by a physician. Assessments included a written questionnaire on lifestyle habits and cardiovascular risk factors, as well as the collection of blood for serum biomarkers and genotyping analysis ^19^. Of 6,889 participants attending the respective exams, 553 had prevalent CVD, 997 had missing clinical components, and 287 had no available genotype data, leaving 4,090 participants with complete LE8 and genetic data (Offspring n=1,255 and Gen3 n=2,835). (**Figure 1)** These exclusions corresponded to participants with prevalent CVD, missing clinical components, or unavailable genotype data. These exclusions corresponded to participants with prevalent CVD, missing clinical components, or unavailable genotype data. De-identified data and materials from the FHS are publicly accessible via the National Institutes of Health Database of Genotypes and Phenotypes. The study was approved by the Boston University Medical Campus Institutional Review Board, and all participants provided written informed consent.

**Figure 1.**
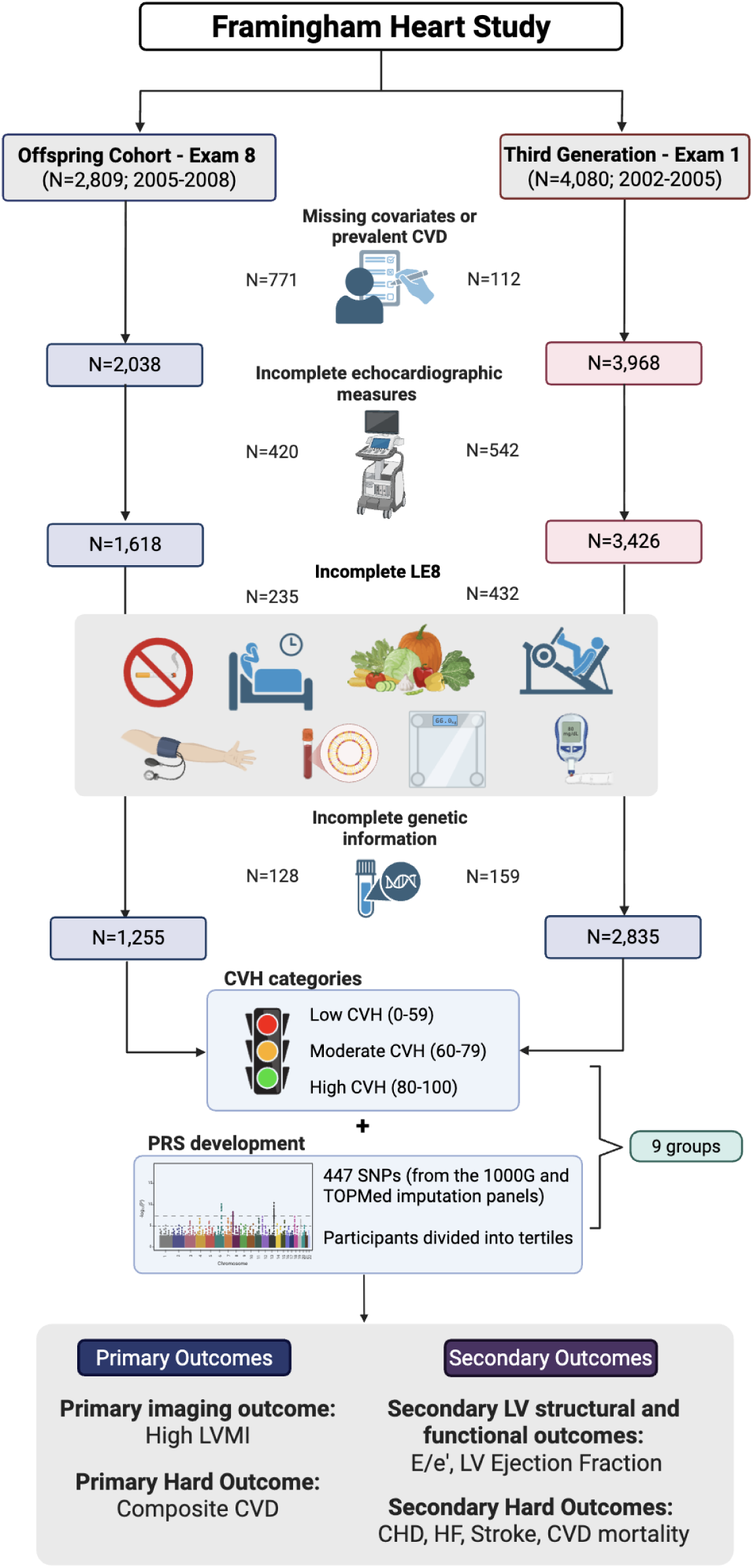
Study flow diagram of the Framingham Heart Study sample selection and outcomes. Participants from the Offspring (Exam 8, 2005–2008) and Third Generation (Exam 1, 2002–2005) were included. After excluding individuals with prevalent CVD, incomplete echocardiography, or missing genetic data, 4,090 participants (1,255 Offspring; 2,835 Gen 3) remained for analysis. Life’s Essential 8 (LE8) cardiovascular health scores were categorized as low, intermediate, or high, and polygenic risk scores (PRS) for LV mass were divided into tertiles, forming nine joint LE8–PRS groups. Created with BioRender.com. *Abbreviations: CVD = cardiovascular disease; LE8 = Life’s Essential 8; LV = left ventricle; PRS = polygenic risk score*.

### Quantification of CVH

The LE8 score was calculated using the guidelines in the report by Lloyd-Jones et al. ^3^, Participants provided self-reported data or FHS examination-measured values. The criteria for diet, physical activity, blood glucose, and blood pressure were modified due to data availability, as validated in previous studies (**Supplemental table)** ^20,21^. The overall LE8 score was calculated for each participant as the unweighted average of the individual component scores, with a score of 80 to 100 considered high CVH, 60 to <80 classified as intermediate CVH, and 0 to <60 as low CVH (20).

### Computation of Polygenic Risk Score for LVMI

To assess genetic predisposition to elevated LVMI, we constructed the PRS for each participant using the published PRS from the referent study (PGS003427) and imputed SNP information from the FHS data ^13^. SNP data were imputed using the 1000G Phase 3, version 5, European samples as a reference panel. Additional SNPs were imputed using the Trans-Omics for Precision Medicine (TOPMED) panel. We retained SNPs with imputation quality greater than 0.4. In total, to compute the PRS for our samples, we used 447 (410 from the 1000G and 37 from the TOPMed imputation panels) out of 465 genetic variants specified in the referent study.

### LV Structure and Systolic and Diastolic Function

During Offspring Exam 8 and Gen 3 Exam 1, participants underwent echocardiography using an HP Sonos 5500 ultrasound system (Philips Medical Systems, Andover, MA) ^22^. 2D echocardiographic images were analyzed offline (EchoPAC, GE Healthcare) using standardized methods to measure linear LV septal and posterior wall thickness and cavity size on parasternal long-axis views. From these measures, LV mass was calculated using the formula LV mass=0.8×[1.04×(EDD+EDST+EDPWT)^3^−(EDD)^3^]+0.6 and indexed to body surface area (BSA) ^23^. LV structure and systolic function were further evaluated using LV ejection fraction (LVEF), calculated using the modified biplane Simpson’s method using cavity measurements in two-chamber and four-chamber views ^24^. As a measure of LV diastolic function, we measured the ratio of early-diastolic peak mitral inflow (E wave) velocity to the averaged peak diastolic lateral and septal mitral annular velocities (e’), labeled E/e’.

### Outcomes of Interest

Incident hard outcomes of interest included CHD, HF, stroke, and a composite CVD, defined as CHD, HF, peripheral artery disease, or stroke. Stroke was a composite event that included transient ischemic attack, ischemic stroke, embolic stroke, and hemorrhagic stroke. Following standardized FHS criteria, CV events were determined by a committee of 3 investigators who reviewed comprehensive inpatient and outpatient medical records.

### Statistical Analysis

Baseline characteristics were summarized as mean (SD) or median (Q1, Q3) for continuous variables and as n (%) for categorical variables. Our predictor variable was the combination of LE8 and PRS, generating 9 distinct groups. We constructed tertiles of LE8 and PRS (each low, intermediate, and high groups), resulting in 9 groups of LE8 x PRS, whereby the optimal risk (high LE8 + low PRS) group served as the referent. We defined a primary imaging outcome of high LVMI (>median value) and a primary hard CV outcome as the composite of CHD, HF, stroke, or Peripheral Artery Disease. Secondary LV structural and functional outcomes included LVMI, LVEF, and E/e’, and secondary outcomes included CHD, stroke, and CVD mortality considered separately.

We used multivariable-adjusted mixed linear regression models and Cox proportional hazards regression models to evaluate the association of LE8 and PRS groups with outcomes mentioned above ^25^. We tested the proportional hazards assumption and applied multivariable-adjusted non-proportional hazards models when the proportional hazards assumption was violated ^25^. We included the following variables in two models 1) age and sex, and 2) Multivariable-adjusted for age, sex, waist circumference, heart rate, C-reactive protein, and creatinine ^26,27^. We evaluated time-varying hazard ratios by evaluating interactions between independent variables and log-transformed time in days. Beta coefficients, standard errors, and p-values were estimated for each outcome to assess their composite group effects.

We evaluated for effect modification by age and sex in the relations of LE8-PRS groups with all outcomes by incorporating corresponding interaction terms (age was coded as 0/1 for ≤ median age vs > median age, respectively). Interaction term p-values <0.05 were deemed statistically significant. If interaction terms were deemed significant, we conducted stratified analyses. All the analyses were run on *R* (version 4.1.2).

## RESULTS

### Baseline Characteristics

The study sample included 4,090 participants from the Offspring (N=1,255) and Gen 3 (N=2,835) cohorts, with a mean age of 47 years (56% female). Characteristics of our study sample are presented in **Table 1**. The mean LE8 score was 70, indicating intermediate CVH. Cardiometabolic markers, including CRP and creatinine levels, were comparable between cohorts. In Gen 3 LE8 scores, including BMI, blood pressure, and glycemic control, were generally higher, consistent with more favorable values (**Table 2**).

**Table 1.**
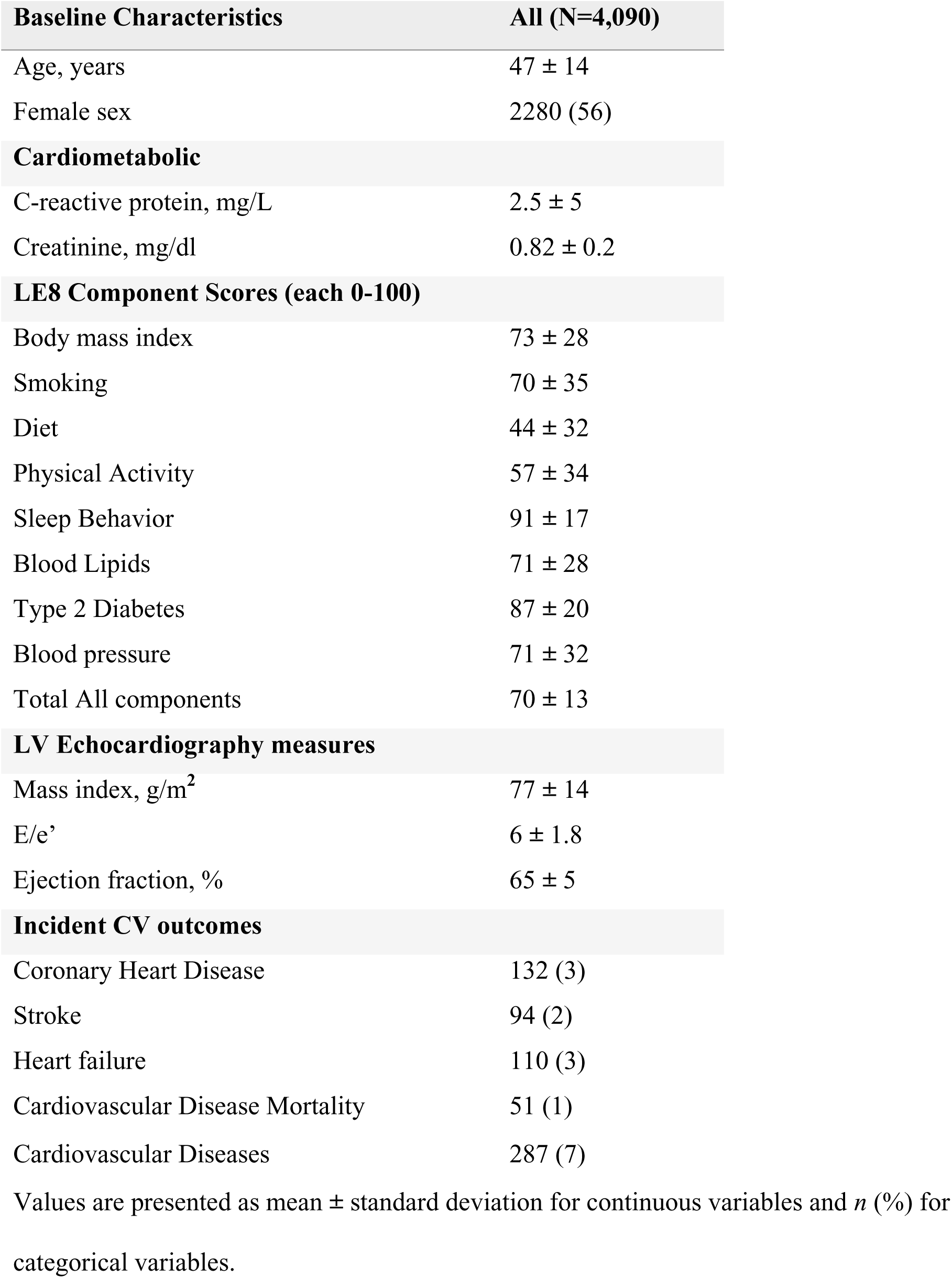
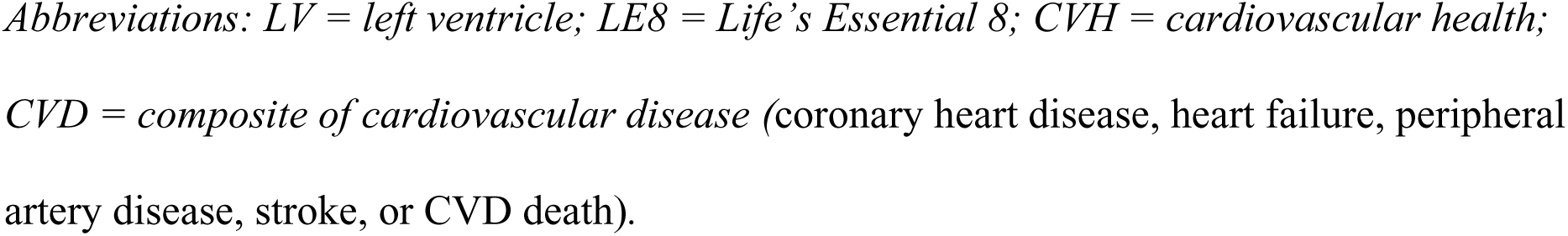
Baseline Characteristics.

**Table 2.**
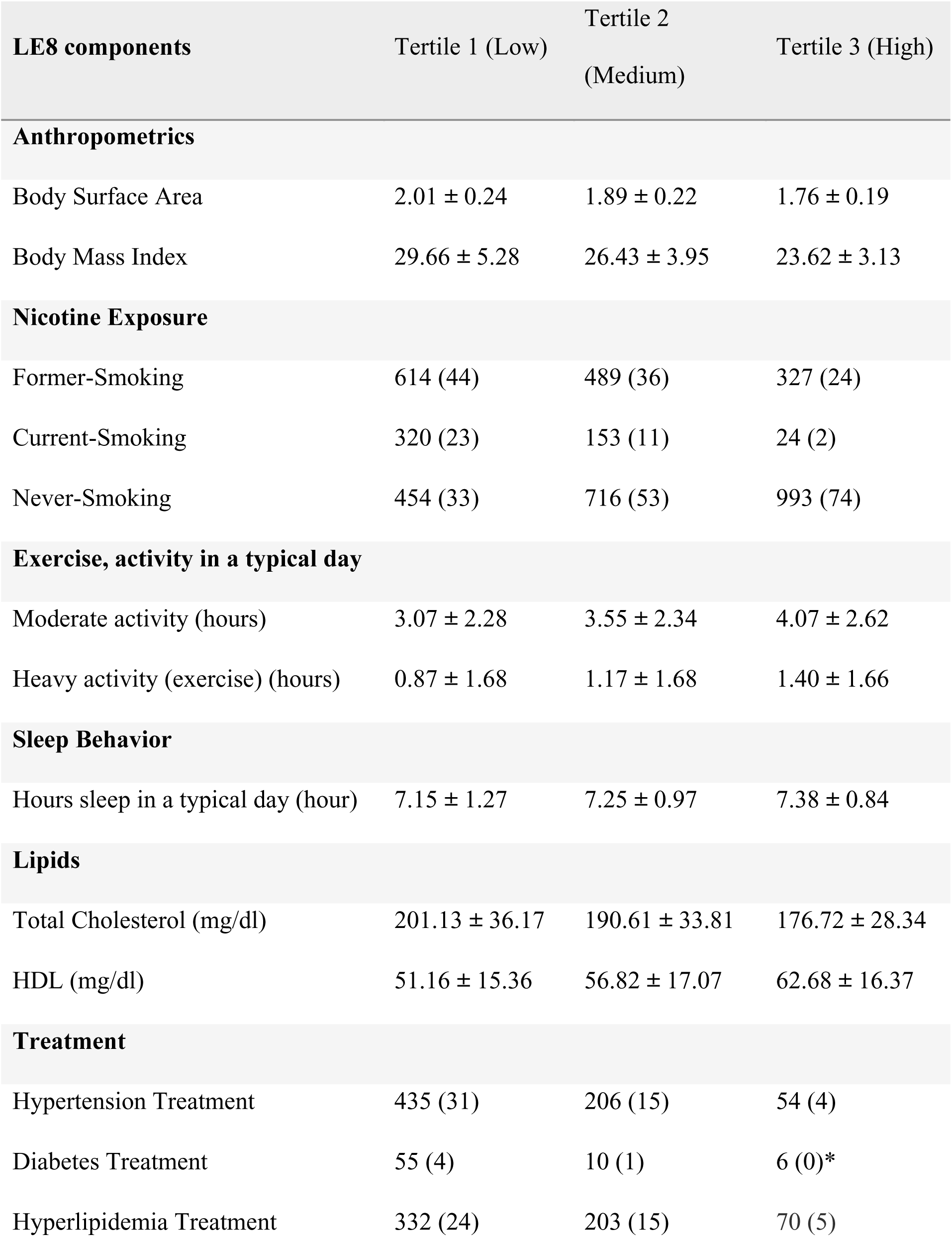
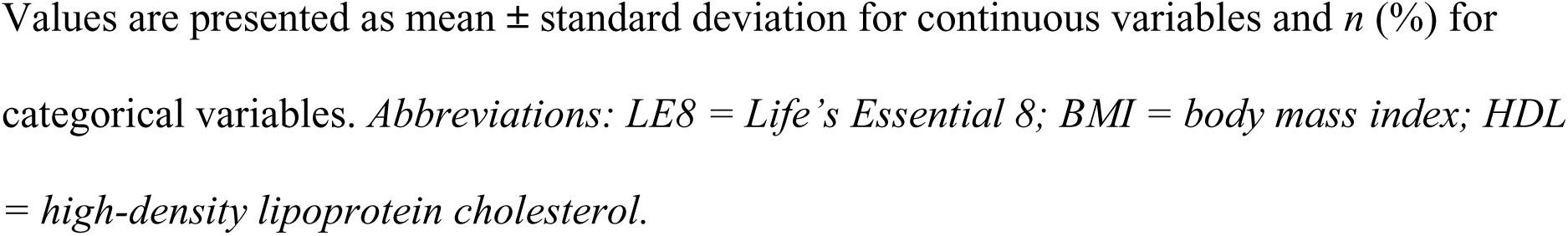
Summary statistics for LE8 components.

### Relations of LE8 and PRS with Cardiac Remodeling and Function

A total of 1,023 participants had high LVMI (defined as ≥75 percentile). Participants in the low LE8-high PRS group had higher values of LVMI compared to the referent **(Table 3 and Figure 3)**. Compared with the referent group (high LE8-low PRS), high PRS was associated with elevated LVMI across tertiles of lower (more unfavorable) LE8 scores, with LVMI values over 2 g/m^2^ in participants with high LE8 to over 4 g/m^2^ in the low LE8 group [multivariable-adjusted β±SE: 4.37±0.80, p<0.0001]. Participants with low LE8 scores, at all levels of PRS, had higher E/e’ values, a measure of LV diastolic dysfunction, with highest effect size in the least favorable LE8-PRS group (multivariable-adjusted β±SE: 0.60±0.10, p<0.0001). We did not observe a significant relation between the LE8-PRS groups with LVEF.

**Table 3.**
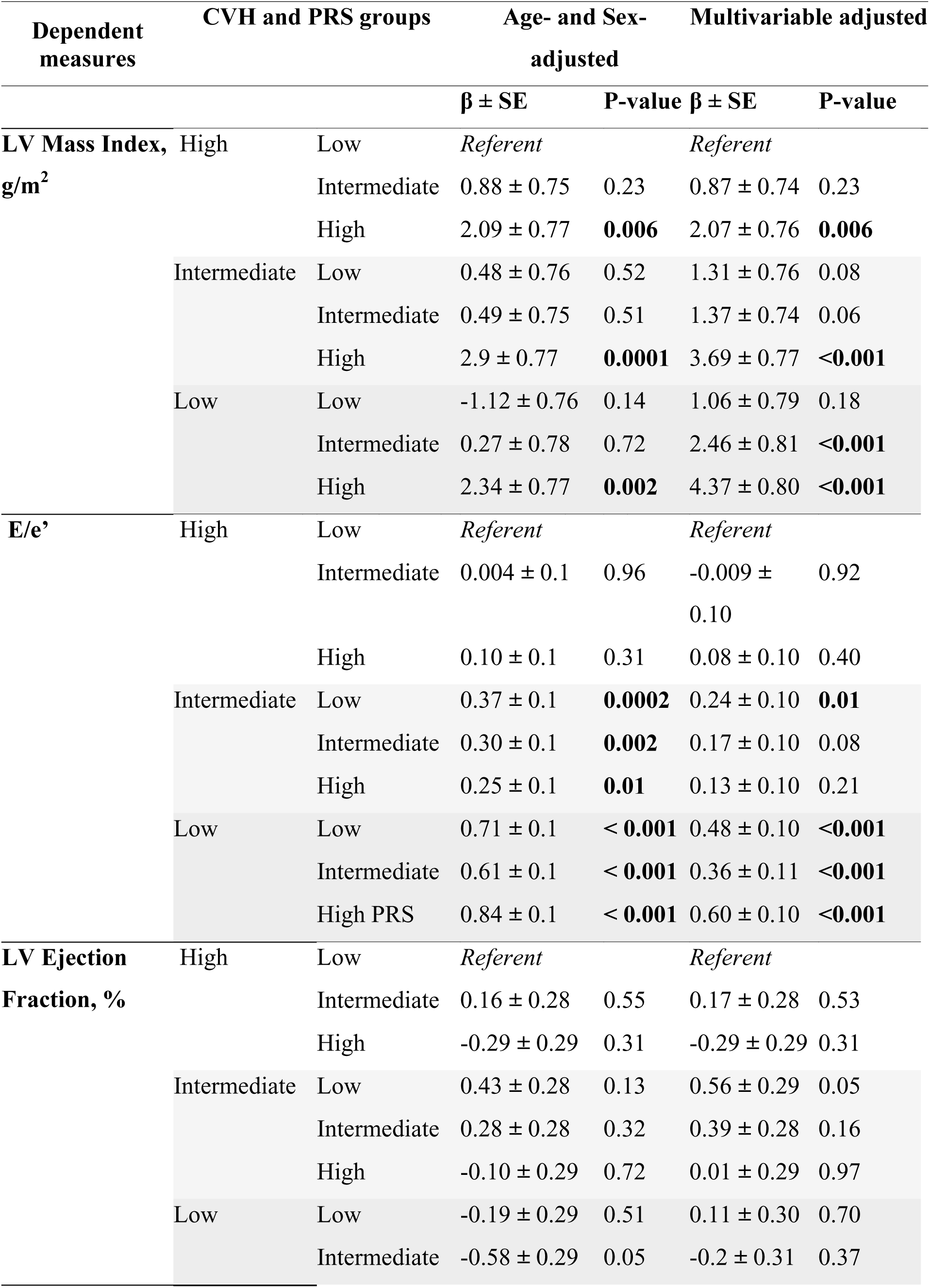
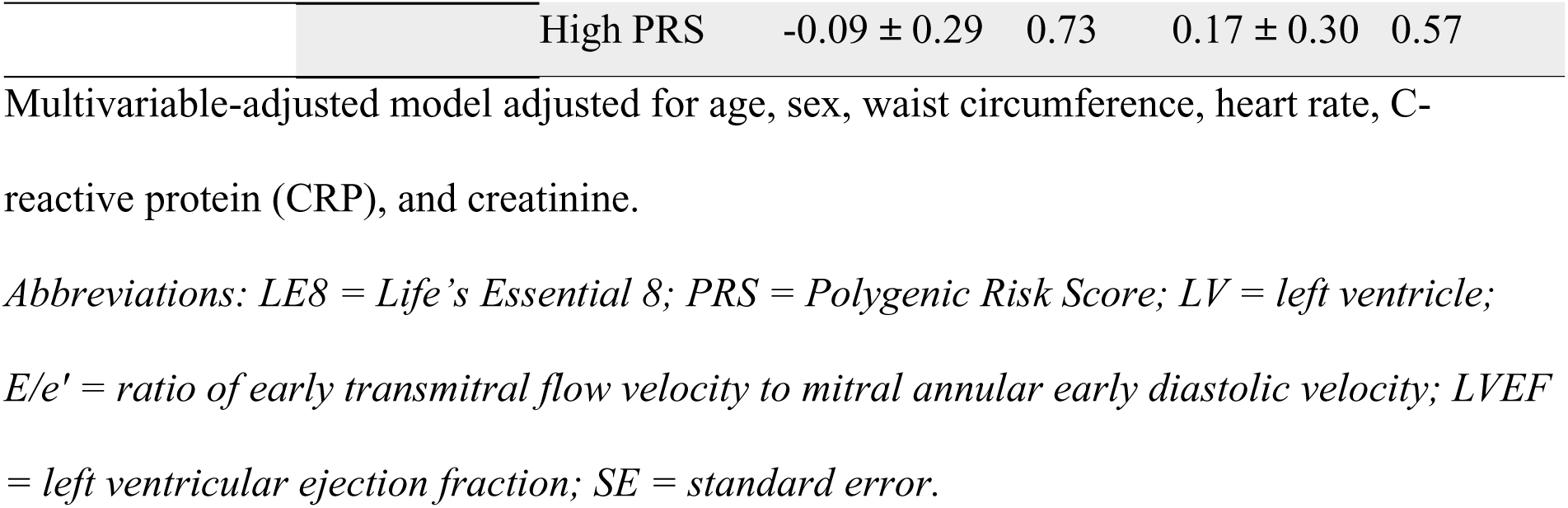
Association of LE8 and PRS with Left Ventricular Structure and Function.

**Table 4.**
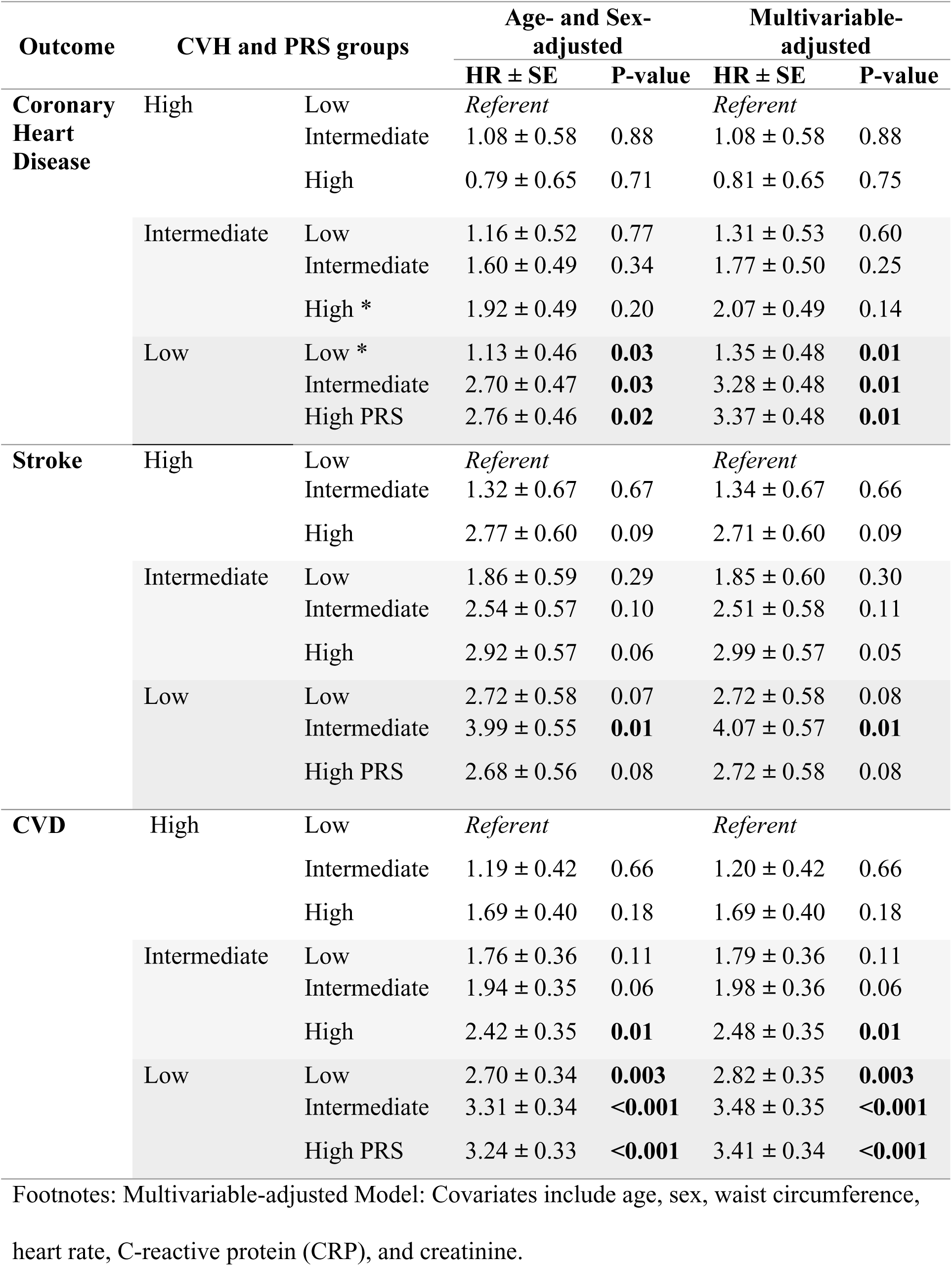
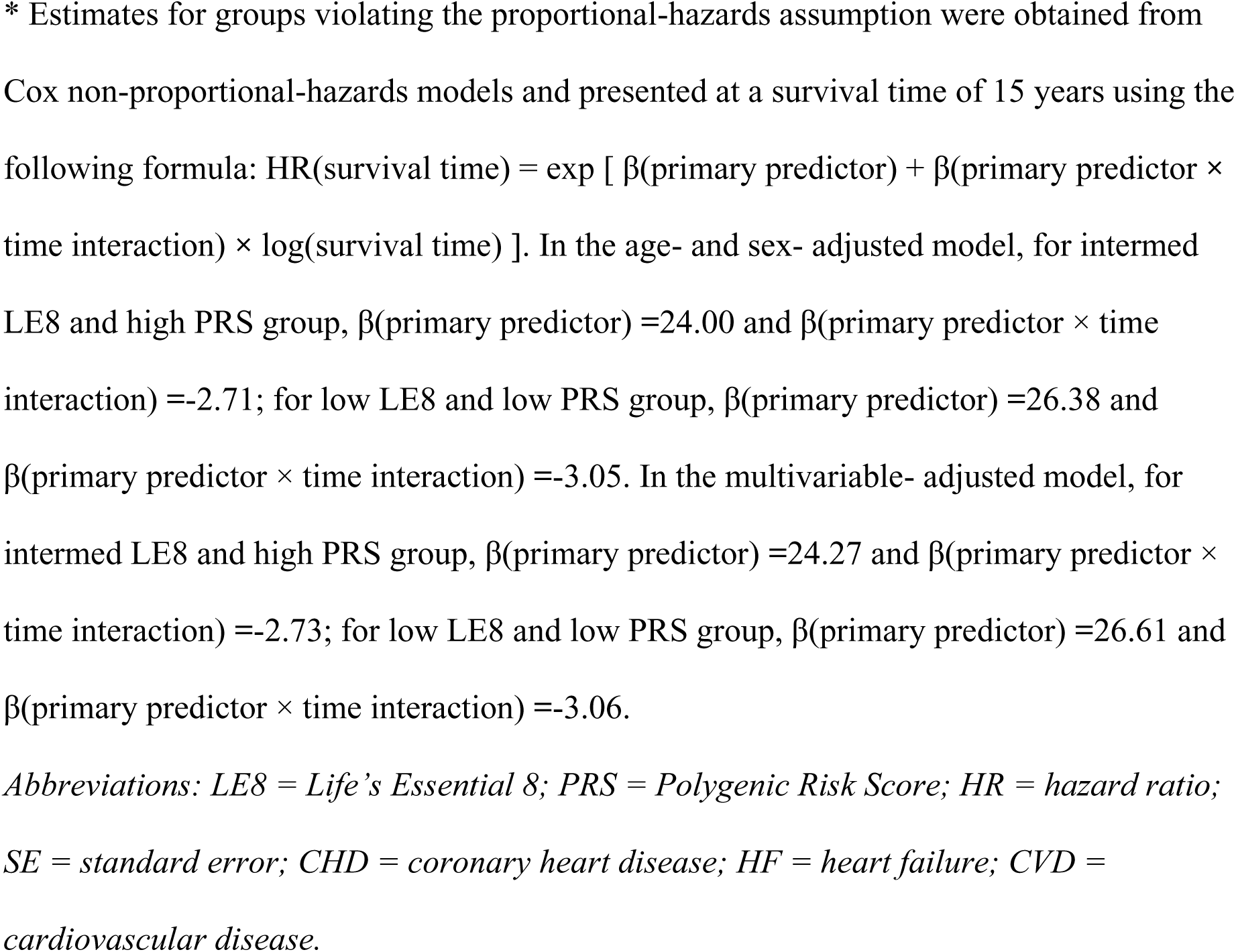
Associations of LE8 and PRS with Incident CV Outcomes.

### Relations of LE8 and PRS with Incident CVD outcomes

The median follow-up time of participants was 19 years (range 0.17-21 years; 15 years in the Offspring and 19 years in Gen 3). We observed 110 incident stroke events, 132 CHD events, and 287 composite CVD events, corresponding to incidence rates of 1.53, 1.84, and 4.06 per 1,000 person-years, respectively. We observed a significant association of the LE8-PRS with CVD development, with participants in the intermediate LE8-high PRS group having an approximate 2.5-fold higher risk compared to those in the referent group **(Table 4 and Figure 3)**. The highest risk for CVD was observed in the low LE8 group, particularly among intermediate and high PRS groups compared with the referent group (multivariable-adjusted HR=3.41±0.34, p<0.001).

Participants having low LE8 scores at all PRS levels were at a higher risk of CHD compared to the referent group. Individuals with a low LE8 score combined with an intermediate to high PRS had the highest risk for incident CHD **(Table 4**, **figure 3)**. In the association of tertiles with CHD, two groups—low LE8 + low PRS and intermediate LE8 + high PRS—violated the proportional hazards assumption. For these, hazard ratios were estimated at a specific survival time point (time=15 years), HR=1.35±0.27; p<0.001 and HR: 2.07±0.34; p<0.001, respectively. Stroke risk was highest in low LE8 + intermediate PRS (HR=.07 ± 0.57, p=0.01) (**Table 4**).

### Effect Modification by Age and Sex in the Relations of LE8 and PRS with Cardiac Remodeling and CVD

We observed significant effect modification by age in the association between LE8 + PRS categories and LVMI (p<0.001). Among participants younger than the median age of 47 years, higher PRS levels were associated with greater LVMI, particularly in those with less favorable cardiovascular health. Compared to the reference group, individuals in this age group with High LE8+High PRS, as well as Low LE8+Intermediate PRS had higher LVMI (β=1.71±0.84, p=0.04; β=2.19±1.04, p*=*0.03, respectively). In contrast, participants older than 47 years of age showed more marked increases in effect size with declining LE8 scores across all PRS strata, with the highest values observed in the Low LE8+High PRS group (β=7.55±1.31, p<0.001) compared to the referent (**Figure 2)**.

**Figure 2.**
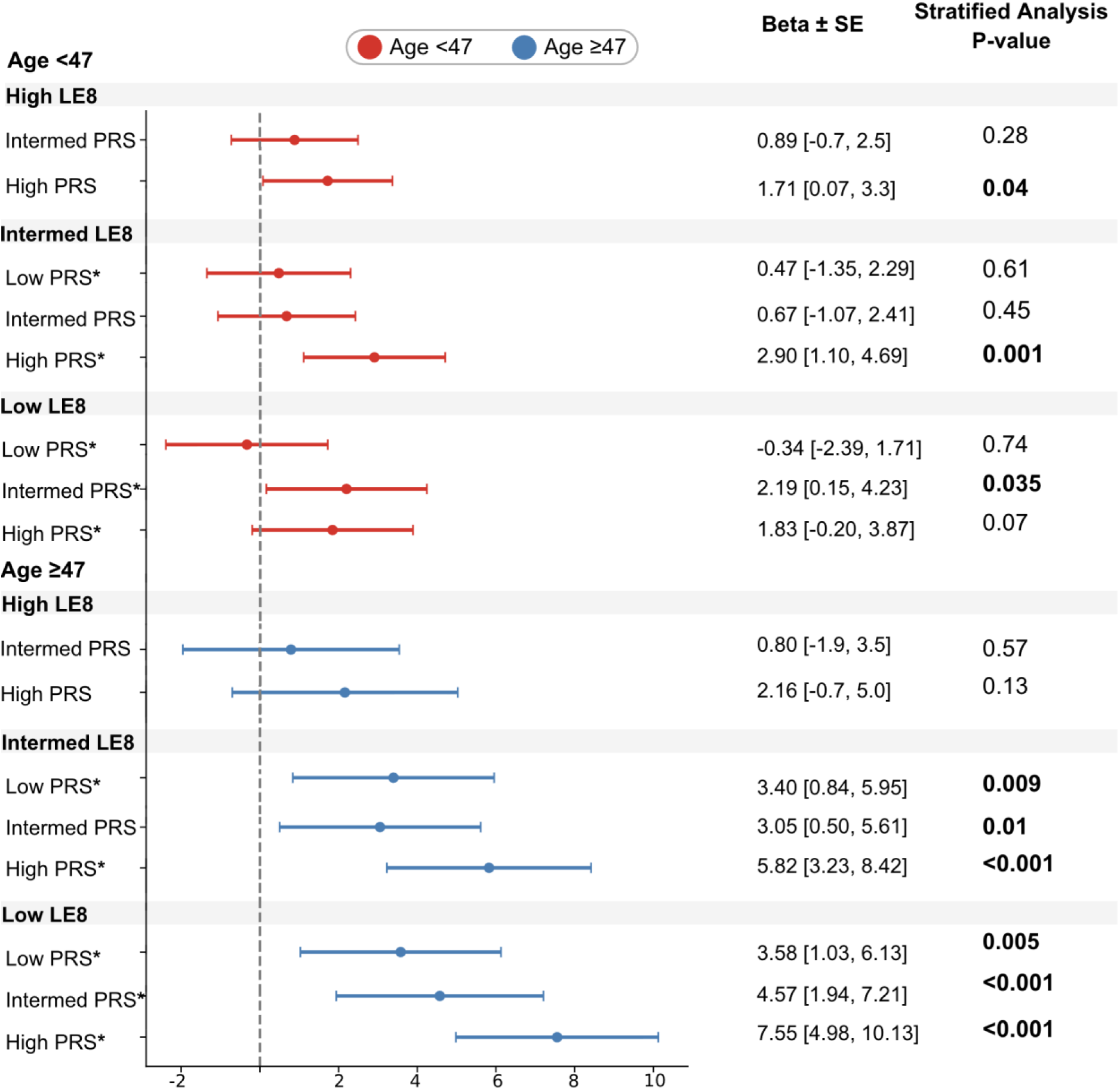
Association of LE8 and PRS with LV mass index in age <47 vs age ≥47 groups in the multivariable-adjusted model. Forest plot showing the associations of Life’s Essential 8 (LE8) cardiovascular health categories and Polygenic Risk Score (PRS) tertiles with left ventricular mass index (LVMI), stratified by median age <47 (red) vs ≥47 years (blue). The X-axis shows beta coefficients and standard errors from the multivariable model; the Y-axis represents combined LE8–PRS groups. We observed a higher effect size in the association between LE8/PRS group with LVMI in individuals ≥47 group, compared to the referent group (High LE8+Low PRS). (*) Denotes age interaction P-value <0.04 *Abbreviations: LE8 = Life’s Essential 8; PRS = Polygenic Risk Score; LVMI = left ventricular mass index*.

**Figure 3.**
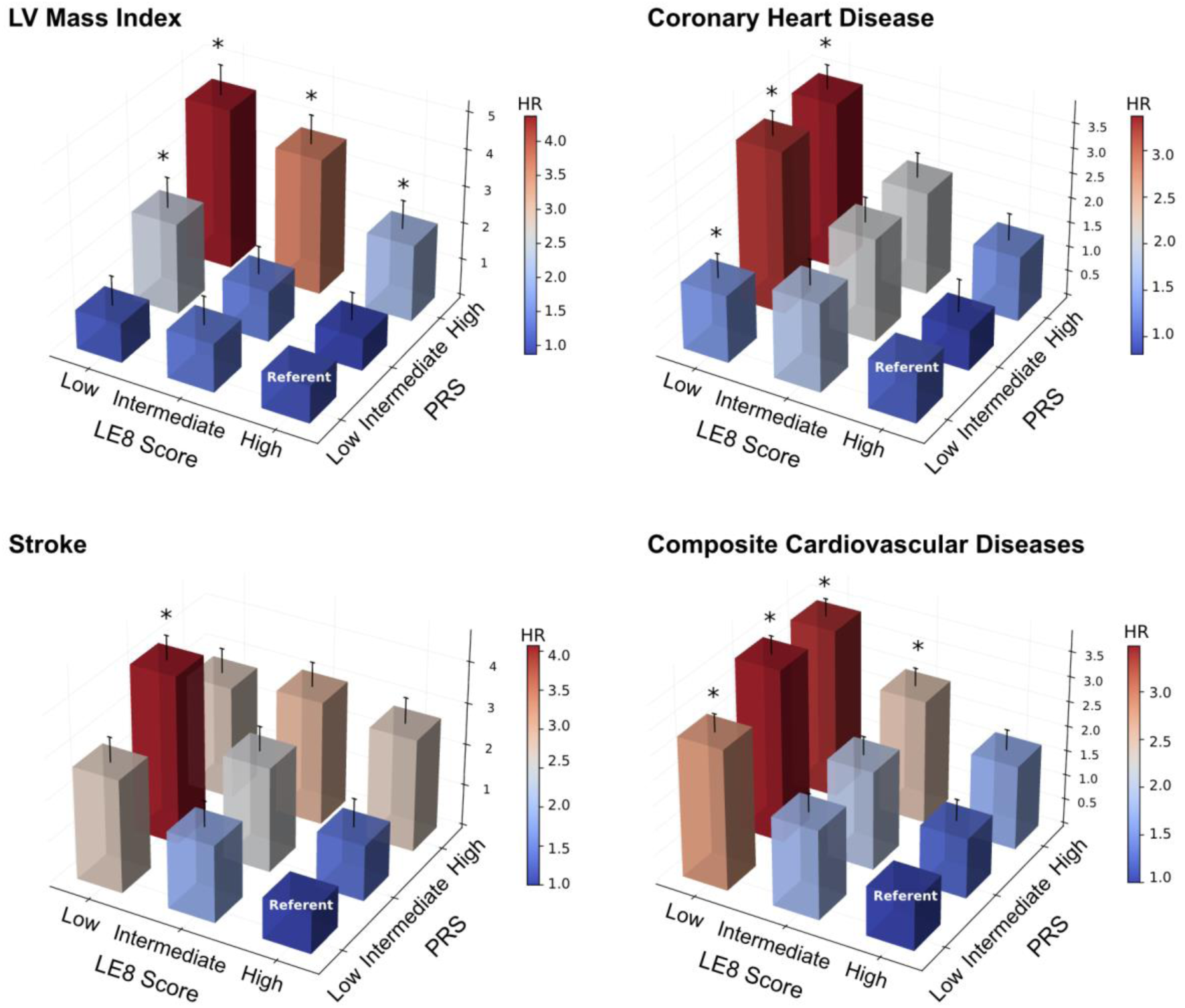
Association of LE8 and PRS with CVD outcomes. Three-dimensional bar plots showing hazard ratios (HRs) for left ventricular (LV) mass index, coronary heart disease (CHD), stroke (composite of Transient Ischemic Attack, Atherothrombotic, Embolic and Hemorrhagic), and composite cardiovascular disease (CVD) outcomes across combined categories of Life’s Essential 8 (LE8) cardiovascular health score and Polygenic Risk Score (PRS). Bars represent HRs from the multivariable-adjusted model (scale adjacent to each figure), and error bars indicate standard errors. Asterisks (*) denote statistically significant associations. The referent group is High LE8 + Low PRS. For CHD, HRs shown for groups that violated the proportional-hazards assumption correspond to the time-specific non–proportional-hazards estimates used in Table 4; all other groups reflect HRs from the proportional-hazards model. *Abbreviations: LE8 = Life’s Essential 8; PRS = Polygenic Risk Score; CHD = coronary heart disease; CVD = cardiovascular disease; LV = left ventricle*.

We also observed a significant sex interaction in the relations between PRS–LE8 groups and E/e’, a measure of LV diastolic dysfunction (*p*=0.003). In multivariable-adjusted models, compared to referent, women in the Low LE8 groups (whether +High or Low PRS) had higher E/e′ values (β=0.84±0.16, *p*<0.001 and β=0.70±0.15, p<0.001, respectively). While low LE8 was also associated with higher E/e’ in men, the magnitude of these associations was lower (Low LE8+Middle PRS: β=0.43±0.16, p=0.008; Low LE8+High PRS: β=0.43±0.16, *p*=0.007).

There was no significant age- or sex-interactions for the relations between LE8+PRS groups and CVD outcomes (all p>0.05).

## DISCUSSION

### Principal Findings

In this study of 4,090 community-dwelling adults of intermediate CVH across a broad age range, we evaluated the joint influence of modifiable lifestyle factors and inherited genetic risk on cardiac remodeling and incident CVD. We observed several key findings. First, both lower LE8 scores and higher LVMI PRS scores were independently and jointly associated with greater LVMI and worse diastolic function, reflected by higher E/e′ ratio. These associations were most pronounced among individuals with Low LE8+High PRS. Second, we identified significant effect modification by age and sex in the relations of LE8-PRS with LV structure and function, whereby the joint association of poor CVH and high genetic risk on LV mass was amplified with advancing age, and the adverse association of low LE8 with diastolic dysfunction was stronger among women than men. Third, participants with less favorable LE8 and higher PRS had a markedly greater risk of coronary heart disease and composite cardiovascular outcomes, with a three-fold greater risk compared with those in the healthiest group.

### Comparison to the literature

Prior epidemiologic studies have demonstrated that unfavorable levels in individual LE8 components and genetic risk are both independently associated with higher LV mass and consequently greater CVD risk ^28–30^. Associations have been reported for dyslipidemia, hyperglycemia, poor diet, physical inactivity, smoking, and inadequate sleep, all of which are individually linked to greater LVMI and higher CVD incidence ^30^. Among LE8 components, body mass index and blood pressure have shown the most robust associations with cardiac remodeling as seen in the Bogalusa Heart Study, Multi-Ethnic Study of Atherosclerosis (MESA) and UK Biobank Imaging Enhancement Study ^31–33^. In parallel, PRS have refined cardiovascular risk prediction by reclassifying individuals into more accurate risk categories ^8,9,34^. However, evidence on the joint influence of CVH and genetic risk on LV structure and function remains limited. We observed the strongest associations for high LVMI in the Low LE8+Intermediate/High PRS categories, supporting a stepwise and additive pattern in which both unfavorable lifestyle and genetic burden contribute jointly to the development of CVD. However, higher CVH was associated with lower LVMI even among individuals with high PRS, indicating the ability of beneficial lifestyle and modifiable risk factors in countering the effects of genetic risk on adverse ventricular remodeling. These findings may be partly explained by the well-described effects of favorable cardiovascular health on reducing hemodynamic load, neurohormonal activation, metabolic stress, and systemic inflammation, mechanisms that can mitigate ventricular remodeling regardless of genetic susceptibility ^31,35^. We observed progressively less favorable LV diastolic function, measured by higher E/e′ ratio, across lower LE8 scores in all PRS strata, with the strongest impairment in the Low LE8–High PRS group. Although no published studies have directly examined the relationship between LE8 and E/e′ ratio in community-based cohorts, mechanistic and epidemiologic data strongly support that risk factor domains measured by LE8 — particularly blood pressure, adiposity, glycemic control, and sleep health —independently and interactively contribute to higher E/e′ ratio, reflecting increased LV filling pressures and subclinical myocardial diastolic dysfunction ^36–39^. Consistent with these findings, longitudinal population-based data indicate that blood pressure and pulse pressure relate to changes in E and A waves and the E/A ratio, diabetes and BMI to worsening e′ velocity and E/e′, and smoking and HDL-cholesterol to alterations in deceleration time, parameters present in impairment of LV relaxation ^40^.

These associations between LE8–PRS groups and LV structure and function varied by age and sex. Among participants younger than 47 years, higher PRS was modestly associated with greater LV mass index, mainly in those with low LE8 scores, suggesting the strong influence of suboptimal risk factors and behaviors, whereas these associations were stronger and more consistent across LE8-PRS strata in older adults, indicating a greater LV remodeling intensified with age. We also observed a sex interaction in the relations between LE8-PRS and E/e’, such that lower LE8 scores were strongly associated with higher E/e′ ratios in women, consistent with worse diastolic function; these effect sizes were weaker and less consistent among men. These findings align with prior evidence showing that women — particularly older women —exhibit higher E/e′ ratios and greater susceptibility to diastolic dysfunction and elevated LV filling pressures ^41^. In the PURSUIT-HFpEF study, elderly women demonstrated significantly higher E/e′-related indices, including afterload-related diastolic elastance, than men, especially among those aged ≥75 years ^42^.

We also demonstrated that the combination of low LE8 scores and high PRS conferred higher risk for CHD and CVD, supporting the role of modifiable risk factors in these outcomes. These findings have been highlighted in previous studies emphasizing the importance of lifestyle interventions in mitigating cardiovascular risk, regardless of genetic predisposition ^30,43^. A clear gradient of risk conferred by poorer CVH (i.e., low LE8 scores) was observed in our primary outcome (composite CVD), whereby those with lower LE8 scores experienced progressively higher risk for incident CVD. Even with favorable PRS, individuals in the Low LE8 group had a 2.8-fold higher risk compared to referent, rising to 3.4-fold with intermediate and high genetic risk groups. Systematic reviews and meta-analyses demonstrate that higher LE8 scores are associated with significantly lower risk of CV mortality ^5,44^.

Participants with Low LE8+Intermediate PRS had the highest risk of stroke. These findings align with prior population studies demonstrating that increased left ventricular mass and hypertrophy are strong, independent predictors of incident stroke, including ischemic and embolic subtypes. In the Atherosclerosis Risk in Communities Study, each 10 g/m² increment in LVMI was associated with a 15% higher stroke risk, independent of conventional risk factors ^45^. Our study extends these observations by suggesting that genetic predisposition to greater LV mass, captured by the LVMI PRS, may contribute to stroke risk, particularly among individuals with poor cardiovascular health.

### Strengths and limitations

Some limitations of our study bear consideration. First, the observational design of our study limits our ability to establish causal relationships of LE8 and PRS with cardiovascular outcomes. Second, lifestyle behaviors were assessed through self-reported measures, which may introduce misclassification bias. Third, the cohort is predominantly European ancestry and largely based in New England, which may limit the generalizability of our findings to more diverse populations. Finally, we did not formally account for multiple testing.

## CONCLUSIONS

In our large community-based study of adults across a range of CVH and genetic risk for elevated LVMI, both less favorable CVH and higher PRS conferred higher risks across multiple domains, including adverse LV mass, diastolic function, and incident CV events. Our findings underscore the importance of considering modifiable CV risk factors and genetic susceptibility when assessing cardiac structure and cardiovascular risk.

## Data Availability

De-identified data and materials from the FHS are publicly accessible via the National Institutes of Health Database of Genotypes and Phenotypes.

## Abbreviations

CHD: Coronary Heart Disease
CVD: Cardiovascular Disease
CVH: Cardiovascular Health
FHS: Framingham Heart Study
LE8: Life’s Essential 8
LV: Left Ventricle
LVMI: Left Ventricular Mass Index
PRS: Polygenic Risk Score

